# INTESTINAL GLUCONEOGENESIS IS DOWNREGULATED IN PAEDIATRIC PATIENTS WITH COELIAC DISEASE

**DOI:** 10.1101/2021.09.08.21262795

**Authors:** Olof Karlson, Henrik Arnell, Audur H. Gudjonsdottir, Daniel Agardh, Åsa Torinsson Naluai

## Abstract

**Objective:** Untreated coeliac disease (CD) patients have increased levels of blood glutamine and a lower duodenal expression of glutaminase (GLS). Intestinal gluconeogenesis (IGN) is a process through which glutamine is turned into glucose in the small intestine, for which GLS is crucial. Animal studies suggest impaired IGN may have long-term effects on metabolic control and be associated with development of type 2 diabetes and non-alcoholic fatty liver disease (NAFLD). The aim of this study was to thoroughly investigate IGN at the gene expression level in children with untreated coeliac disease.

**Design:** Quantitative polymerase chain reaction (qPCR) was used to quantify expression of 11 target genes related to IGN using the delta-delta Ct method with three reference genes (*GUSB, IPO8* and *YWHAZ*) in duodenal biopsies collected from 84 children with untreated coeliac disease and 58 disease controls.

**Results:** Significantly lower expression of nine target genes involved in IGN was seen in duodenal biopsies from CD patients compared with controls: *FBP1, G6PC, GLS, GPT1, PCK1, PPARGC1A, SLC2A2, SLC5A1*, and *SLC6A19*. No significant differences in expression were seen for *G6PC3* and *GOT1*.

**Conclusion:** Children with untreated coeliac disease have lower expression of genes important for IGN. Further studies are warranted to disentangle whether this is a consequence of intestinal inflammation or due to an impaired metabolic pathway shared with other chronic metabolic diseases. Impaired IGN could be a mechanism behind the increased risk of NAFLD seen in CD patients.

**SIGNIFICANCE OF THIS STUDY:** *What is already known about this subject?:*

- Genome-wide association studies have shown an association between coeliac disease (CD) and glutaminase (*GLS)*.
- Intestinal gluconeogenesis (IGN) is a process with a recently described important function in energy homeostasis and metabolic disease. *GLS* is critical for IGN by enabling it to use glutamine, its main substrate.
- CD patients are at an increased risk of non-alcoholic fatty liver disease (NAFLD) as adults.

*What are the new findings?:*

- Nine genes involved in IGN are downregulated at the gene expression level in the small intestine of children with untreated CD, suggesting impairment of IGN.

*How might it impact on clinical practice in the foreseeable future?:*

- Impaired IGN might be a mechanism behind the increased risk of NAFLD seen in CD patients as adults.
- Early diagnosis and treatment of CD may restore IGN and prevent CD patients from NAFLD later in adulthood.

## INTRODUCTION

A genome-wide association study (GWAS) recently found shared genes between coeliac disease (CD) and type 2 diabetes (T2D), indicating changes in common nutrient signalling pathways of amino acid metabolism. Glutaminase (GLS), an enzyme that converts glutamine to glutamate, was identified by pathway analysis in the most significant network of genes and was found to be downregulated in duodenal biopsies from children with untreated CD.^1^ Later work showed allele specific expression of *GLS* connected to the associated single-nucleotide polymorphism (SNP) (rs6741418) in the *GLS* gene region.^2^ A total of 142 associations to 50 traits, a large number of which are autoimmune diseases, have been reported for the gene region containing *GLS/STAT1/STAT4* (2021-07-18; GWAS Catalog: https://www.ebi.ac.uk/gwas). This makes it one of the top-reported gene regions in autoimmune diseases so far. Out of these 142 reported associations, 26 SNP variants, have been shown to have a significant influence on the expression of *GLS*, three SNP variants influence the expression of *STAT4*, and none have been shown to influence *STAT1* expression (https://www.gtexportal.org). Supplementary table 1 shows the most associated variant reported for each trait (data from the GWAS Catalog). Thus, there is data indicating a possible common genetic mechanism behind autoimmunity and differential expression of *GLS*. Interestingly, patients with untreated CD also have altered blood levels of several amino acids, including glutamine/glutamate, further suggesting alterations of metabolism warranting more investigation.^3^ Figure 1 illustrates a study flow chart and outline of previous work as well as the work in the present study.

**Figure 1.**
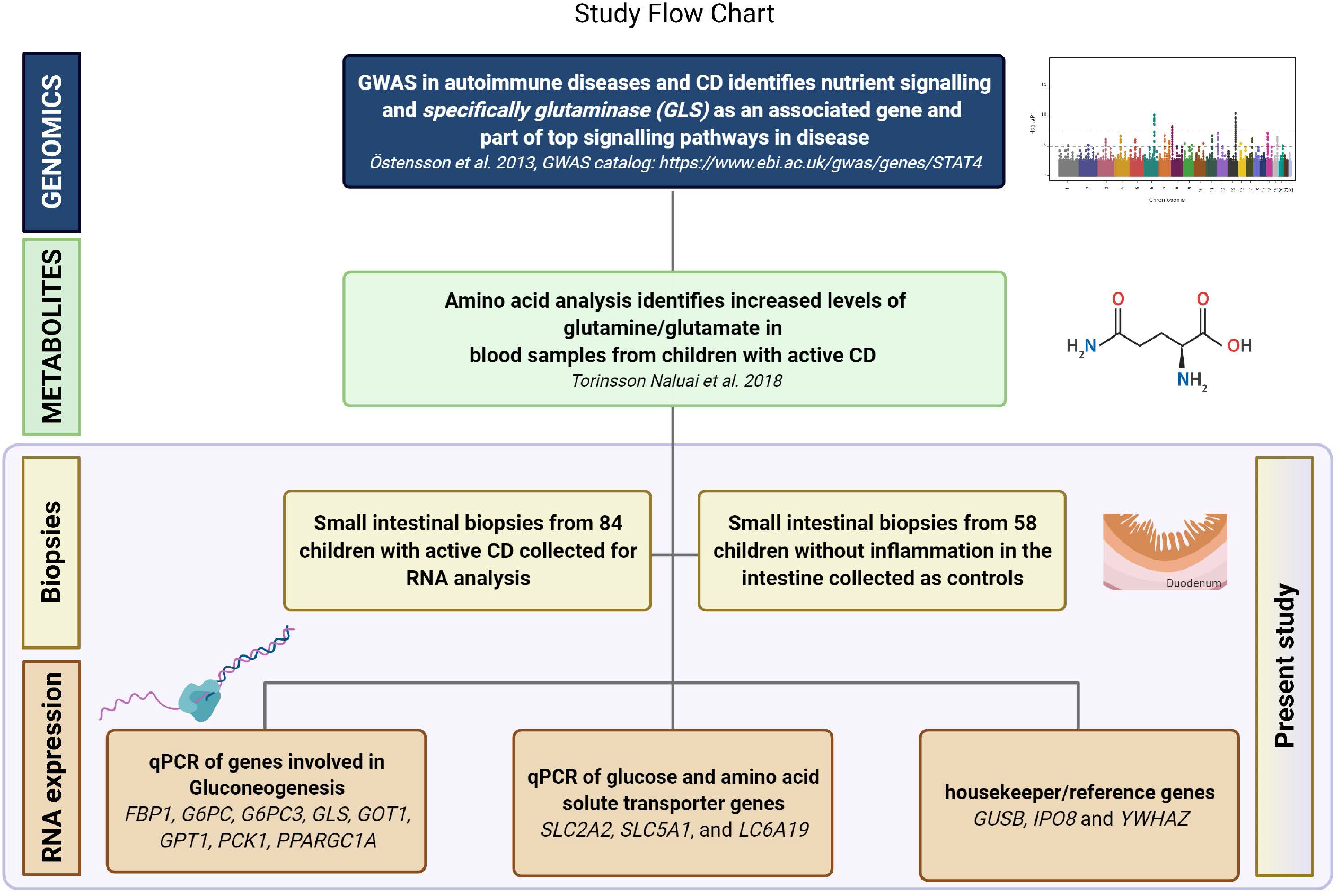
Study flow chart and outline of previous work as well as the work in the present study.

Intestinal gluconeogenesis (IGN) is a process from which glucose can be produced in the small intestine.^4^ Glutaminase has a critical role in IGN since it catalyses the first step needed to use its main substrate, which is glutamine.^5^ In animal models, IGN-deficient mice develop hyperglycaemia.^6^ Mice with induced high levels of IGN instead seem protected against hyperglycaemia, even when fed a high-fat/high-sucrose (HF-HS) diet. IGN-deficient mice also more easily develop hepatic steatosis, both on an HF-HS diet as well as a standard diet, while mice with induced IGN are again protected, even on an HF-HS diet.^7^ In a rat model investigating mechanisms behind bariatric surgery as treatment of T2D, rats showed increased IGN after surgery.^8^ In human studies, T2D patients with high expression of genes involved in IGN had greater improvement of insulin resistance scores after bariatric surgery.^9^ These data indicate an important role for IGN in energy homeostasis and metabolic disease, which is suggested to be explained by a glucose-sensing mechanism in the portal vein, where higher levels of glucose indicating an adequate glucose supply, lead to satiety signals sent via spinal nerves to the brain which then affects whole-body metabolism.^10^

The aim of the present study was to investigate IGN in CD patients by investigating intestinal gene expression of *GLS* and another 10 selected genes involved in IGN in children with untreated CD. Figure 2 illustrates IGN and the roles of the selected genes. The hypothesis was that with *GLS* downregulated, the whole process of IGN with its important metabolic function could be affected.

**Figure 2.**
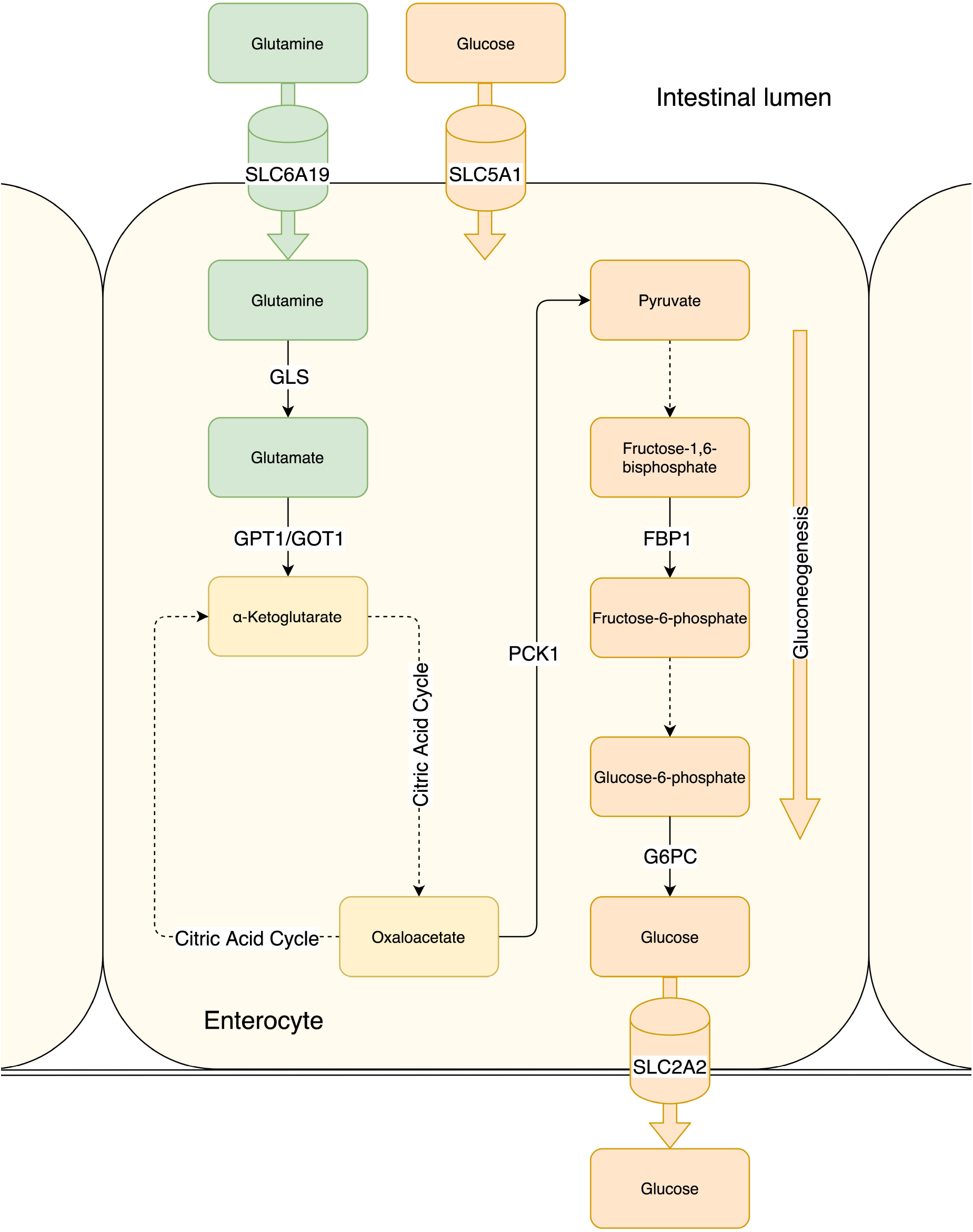
Gluconeogenesis and steps for using glutamine as substrate for gluconeogenesis showing the enzymes for which we have quantified gene expression. Dotted lines represent steps that have been left out of the figure where gene expression has not been investigated. Investigated solute carriers also shown.

## MATERIALS AND METHODS

### Biological material

Duodenal biopsies were collected from children at four different hospitals in Sweden as previously described.^11^ All children had been referred for an upper endoscopy for medical reasons and were consecutively recruited to the study. For this study, 84 untreated CD cases and 58 disease controls were used. Among these, 64 out of 84 (76%) cases and 34 out of 58 (59%) controls, were female. Age of cases was 6.1 ± 3.7 and age of controls was 11.6 ± 4.5 years old (mean ± standard deviation). All disease controls had normal mucosal findings (Marsh score 0-1) and children with inflammatory bowel disease or Helicobacter pylori infections were excluded prior to analysis. Duodenal biopsies were immediately put in RNA-stabilising RNAlater solution (Life Technologies, CA, USA) and put in room temperature overnight to allow the reagent to penetrate the sample. Biopsies were then frozen in -80°C until RNA extraction was performed. Total RNA was extracted using the AllPrep® DNA/RNA/Protein Mini Kit (Qiagen, Germany). RNA quality and quantity were measured with a NanoDrop 2000c spectrophotometer (Thermo Fisher Scientific, MA, USA) and a 2100 Bioanalyzer (Agilent Technologies, CA, USA). RNA was converted to cDNA for storage using the SuperScript Vilo cDNA synthesis kit (Thermo Fisher Scientific, MA, USA) and stored at -80°C.

### Quantitative polymerase chain reaction

Quantitative polymerase chain reaction (qPCR) was performed using TaqMan^®^ technology (Thermo Fisher Scientific, MA, USA). The final reaction consisted of TaqMan^®^ gene primers and equal parts sample cDNA and TaqMan^®^ Universal Master Mix II. All primer-sample reactions were run in duplicate in 384-well plates on a QuantStudio 12K Flex Sequence Detection System (Thermo Fisher Scientific, MA, USA). Target genes were *FBP1* (fructose-bisphosphatase 1), *G6PC* (glucose-6-phosphatase), *G6PC3, GLS, GOT1* (glutamic-oxaloacetic transaminase 1), *GPT1* (glutamic--pyruvic transaminase), *PCK1* (phosphoenolpyruvate carboxykinase 1), *PPARGC1A* (PPARG coactivator 1 alpha), *SLC2A2* (solute carrier family 2 member 2), *SLC5A1* (solute carrier family 5 member 1), and *SLC6A19* (solute carrier family 6 member 19). Reference genes were *GUSB* (glucuronidase beta), *IPO8* (importin 8) and *YWHAZ* (tyrosine 3-monooxygenase/tryptophan 5-monooxygenase activation protein zeta). These reference genes have been evaluated by the group previously using the *Normfinder* software.^12^

### Statistical analysis

Quality control of qPCR data was done in the ExpressionSuite v1.1 program (Thermo Fisher Scientific, MA, USA) where reactions that had not run properly were filtered out. ΔC_T_ values were calculated using R^13^ with the RStudio developing environment (which was used for all work in R)^14^. Group differences were examined by a generalised linear model using the glm function in R, running one model based only on CD case status and one model adjusting for age and sex as covariates. A generalised linear model was used to enable adjustment for both a quantitative (age) and a qualitative variable (sex). With α = 0.05 correcting for multiple testing of 11 target genes using the Bonferroni method gave 0.05/11=0.0045 as the adjusted significance threshold. Data visualisation was done using the ggplot2 package in R,^15^ part of tidyverse^16^ from which other packages were also used when coding in R. Fold change was calculated using the 2^-ΔΔC^_T_ method.^17^ A partial correlation analysis using gene expression levels, tissue transglutaminase (tTG) antibody levels and Marsh score controlling for age was performed in IBM SPSS statistics software version 27. P-values were set as 2-sided. Marsh scores of all cases were transformed to numbers; Marsh stage 2 – 1, Marsh stage 3A – 2, Marsh stage 3B – 3 and finally Marsh stage 3C – 4.

We used the STROBE case-control reporting guidelines when writing this paper (von Elm E, Altman DG, Egger M, Pocock SJ, Gotzsche PC, Vandenbroucke JP. The Strengthening the Reporting of Observational Studies in Epidemiology (STROBE) Statement: guidelines for reporting observational studies).

## RESULTS

Analysing gene expression levels in duodenal biopsies using qPCR, *FBP1, G6PC, GLS, GPT1, PCK1, PPARGC1A, SLC2A2, SLC5A1*, a d *SLC6A 9* showed significantly lower expression in CD cases compared with controls, whereas *6PC3* and *OT1* showed no differences (table 1 and figure 3). This remained when adjusting for age and sex of study participants.

**Table 1.** Gene expression levels in duodenal biopsies from children with untreated coeliac disease (CD) (n = 84) and disease controls (Control) (n = 58) determined by qPCR.

| Gene | p-value | p-value<br>(adjusted for<br>age and sex) | CD Mean<br>ΔCT | Control<br>Mean ΔCT | ΔΔCT | Fold<br>Change | % Change |
| --- | --- | --- | --- | --- | --- | --- | --- |
| *FBP1* | **5.68E-11** | **9.96E-09** | 1.95 | 2.96 | -1.02 | 0.49 | -51 |
| *G6PC* | **1.24E-10** | **2.40E-07** | -1.70 | 0.12 | -1.82 | 0.28 | -72 |
| *G6PC3* | 0.520 | 0.926 | -0.62 | -0.55 | -0.07 | 0.95 | -5 |
| *GLS* | **7.02E-12** | **3.05E-09** | -0.87 | -0.11 | -0.76 | 0.59 | -41 |
| *GOT1* | 0.249 | 0.823 | 0.82 | 0.93 | -0.10 | 0.93 | -7 |
| *GPT1* | **2.08E-04** | **2.81E-03** | -1.25 | -0.69 | -0.56 | 0.68 | -32 |
| *PCK1* | **1.08E-16** | **7.27E-12** | 0.66 | 2.82 | -2.16 | 0.22 | -78 |
| *PPARGC1A* | **1.06E-11** | **1.84E-06** | -3.72 | -2.63 | -1.09 | 0.47 | -53 |
| *SLC2A2* | **1.38E-10** | **3.30E-08** | -0.15 | 0.77 | -0.93 | 0.53 | -47 |
| *SLC5A1* | **1.34E-11** | **4.95E-08** | 2.53 | 3.44 | -0.91 | 0.53 | -47 |
| *SLC6A19* | **5.16E-07** | **8.09E-05** | 1.38 | 2.20 | -0.82 | 0.57 | -43 |

Bonferroni-adjusted significance level for eleven target genes was 0.0045. Significant p-values for target genes are highlighted in bold. The far-right column shows ΔΔCT converted to percentage change in gene expression for CD patients compared to controls.

**Figure 3.**
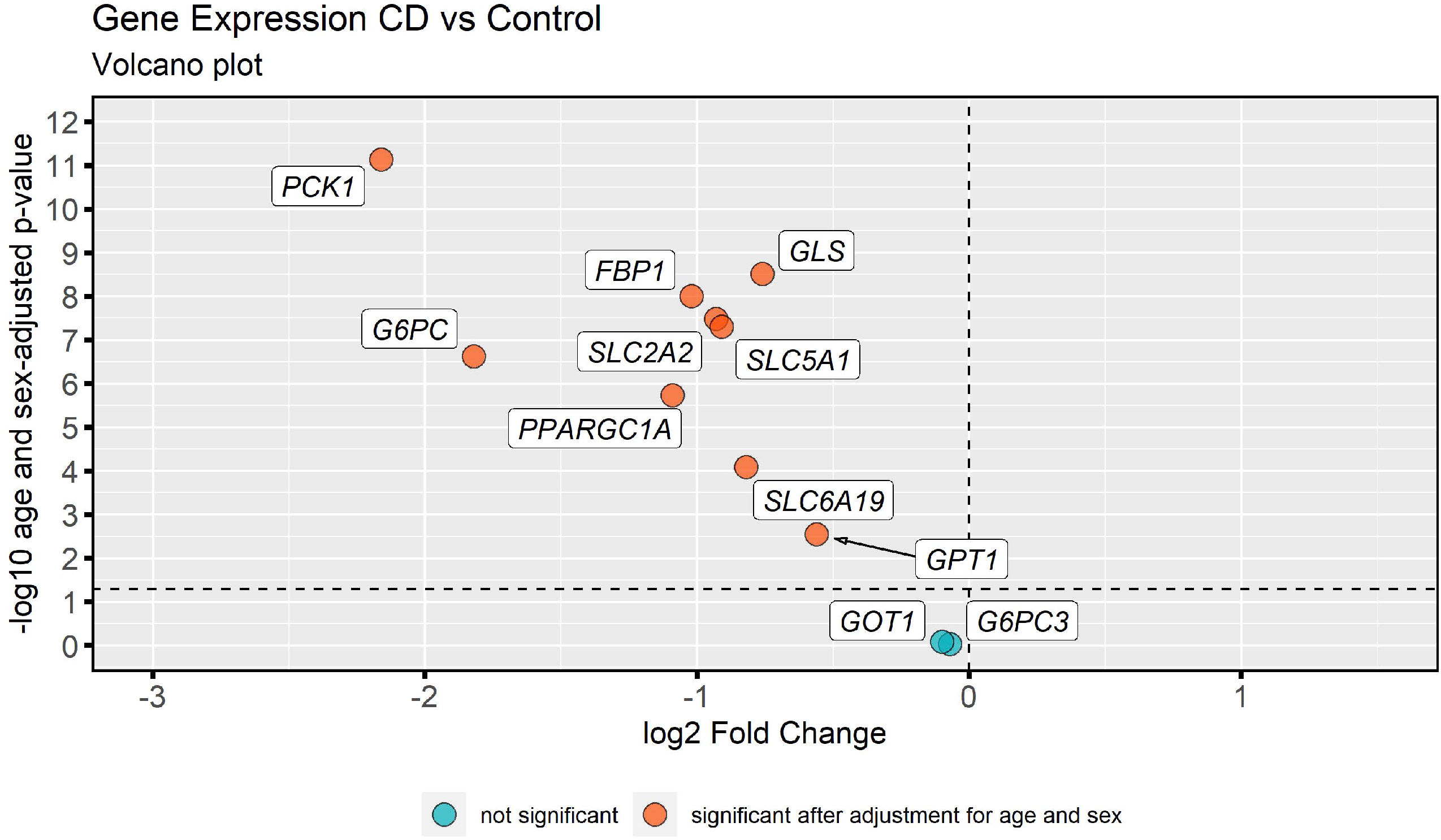
a) Volcano plot showing foldchange on the X-axis and significance (-log p-value) on the y-axis. b) Box plots of gene expression levels in duodenal biopsies from children with untreated coeliac disease (CD) (n = 84) and disease controls (CONTROL) (n = 58). All cases are visualised by individual dots. The middle horizontal line represents the median value, the two “hinges” the first and third quartiles. The “whiskers” extend to the minimum and maximum values within 1.5 interquartile range of hinges. Significance of target genes marked with * P < 0.0045, ** P < 0.00091, *** P < 0.000091 (Bonferroni-adjusted significance thresholds).

Partial correlation analysis in the CD cases (table 2) showed a strong correlation between expression levels of the same nine genes and Marsh scores of patients. *PPARGC1A* also showed a significant correlation to IgA tTG antibody levels, while *FBP1, G6PC, GLS, GPT1, PCK1, PPARGC1A*, and *SLC5A1* showed significant correlation to IgG tTG levels.

**Table 2.**
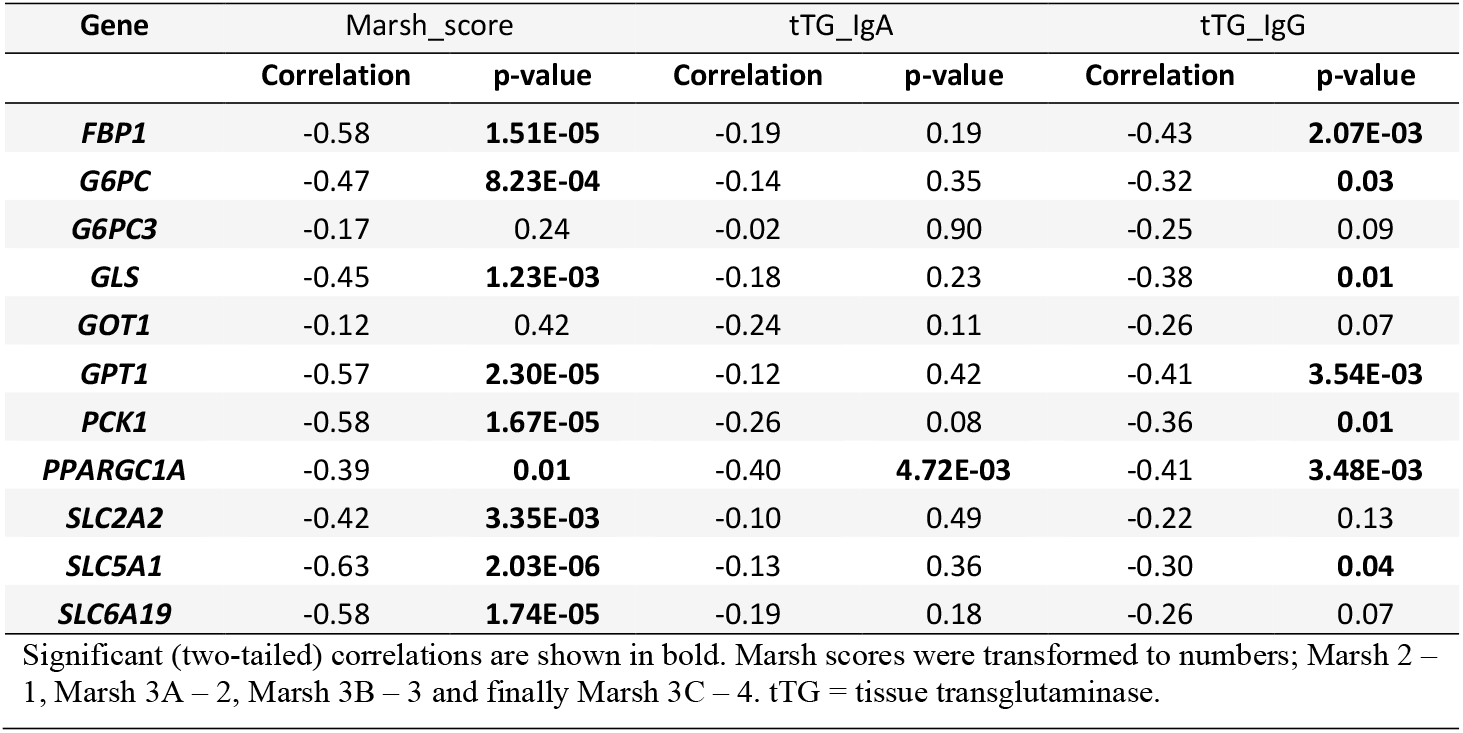
Partial Correlations analysed in cases with CD, controlling for age.

## DISCUSSION

The present study shows that children with untreated CD have a strikingly lower expression of genes involved in IGN in duodenal biopsies when compared with children with a normal intestinal mucosa. Decreased expression is correlated to a higher Marsh score, and to a lesser degree, tTG antibody levels. These results suggest that CD patients could have an impaired function of IGN, either as consequence of chronic intestinal inflammation in untreated disease, but perhaps more interestingly, due to an impaired metabolic pathway shared with other chronic metabolic diseases suggested by the large number of traits shown to be associated with the *GLS*/*STAT1*/*STAT4* gene region. In the long-term perspective, a lower expression of these genes, which are central for IGN and thus important for metabolic homeostasis, may lead to other chronic diseases like NAFLD and T1D if not recognised.

This study provides the first evidence of downregulation of intestinal expression of *G6PC, GPT1, SLC6A19* and *PPARGC1A* in CD. Glucose-6-phosphatase, the protein product of *G6PC*, has an essential role in gluconeogenesis by converting glucose-6-phosphate, which cannot be transported out of the cell, into glucose, which can then be released into the bloodstream, making its downregulation an especially important finding. Decreased expression of *PCK1, FBP1* and solute carriers has previously been shown in adult CD patients, but until now has not been put in the context of IGN.^18-20^ Decreased expression of *GLS* is consistent with our previous finding of its downregulation in the GENEX material.^1^ *G6PC3* and *GOT1* showed no significant changes in expression, however *G6PC* and *GPT1*, which have very similar functions respectively in IGN, are more critical for these respective functions.^21 22^

These data thus imply that the ability of the small intestine to perform gluconeogenesis and release glucose from the intestinal enterocyte might be severely decreased in patients with untreated CD. Decreased expression of *GLS, GPT1, PCK1, FBP1* and *G6PC* suggest an impairment of the IGN pathway from the start of using glutamine in gluconeogenesis, to releasing glucose into the blood. If only *GLS, GPT1* and *PCK1*, were downregulated, using glycerol, the second most important substrate of IGN^5^ could still be possible since it enters gluconeogenesis in later steps, but since *FBP1* and *G6PC*, the protein products of which catalyse crucial final steps in gluconeogenesis, are also downregulated, it stands to reason that the whole pathway of IGN is impaired. We propose that the decreased expression of *PPARGC1A* might provide an explanation for this at a regulatory level since its protein has a key role in regulating hepatic gluconeogenesis. Even though it is unknown if it has a similar role in the intestine, it does not seem entirely implausible. Decreased expression of *SLC6A19* suggests the ability to absorb glutamine, as well as other neutral amino acids transported by the SLC6A19 protein (also known as B0AT1), from the intestinal lumen is impaired. Lower expression of *SLC5A1* (SGLT1) and *SLC2A2* (GLUT2) indicate decreased capacity for glucose transport.

The metabolic effects of impaired IGN in humans are not entirely clear. Studies on IGN in animal models show that an increase in glucose levels in the portal vein provides signals that increase satiety and improve energy homeostasis. Induced high levels of IGN appears to offer protection against metabolic disease, while impairment leads to signs of dysregulated glucose control and hepatic steatosis.^7 10^

CD patients are at increased risk of NAFLD, with the highest risk seen during the first years after diagnosis and the largest relative risk increase seen in patients with a normal BMI.^23 24^ We speculate that impaired IGN could provide an explanation for the increased risk of NAFLD in CD patients. Our study does not examine whether expression of IGN-related genes return to normal in CD patients treated with a gluten-free diet, but the correlation with Marsh scores suggests a lower degree of inflammation might improve IGN. If IGN is normalised when CD is treated, perhaps this could be part of the explanation for why risk of NAFLD is at its highest in the first year after CD diagnosis, when the gluconeogenetic capability of the intestine perhaps has not fully recovered. Such a recovery might also be suggested by a 1968 study of glutaminase enzymatic activity, which found lower levels in untreated CD patients that seemed to recover in patients on treatment with a gluten-free diet.^25^ Studying expression of IGN-related genes in patients before and after treatment would be an important next step.

This study has several limitations. The expression of the selected genes in CD cases were compared with disease controls, i.e., these were children referred for an upper endoscopy investigated for other intestinal diseases affecting the gut. It cannot be excluded that the disease controls may have had conditions that can affect the expression of the selected genes. However, all disease controls had normal mucosal findings and children with inflammatory bowel disease and *Helicobacter pylori* infections were excluded prior to analysis. Another limitation was that cases and controls were not age and sex matched. Still, when adjusting for age and sex the results remained significant. The strength of the study is that children were enrolled from three sites by paediatric gastroenterologists with long clinical experience of diagnosing and treating children with CD. Moreover, all intestinal biopsies were blindly reviewed and scored histologically by a single pathologist before analysis was performed in this study in order to minimise misclassification of cases and controls.

The results from the present study raises several questions. It is not clear whether the downregulation of the target genes is specific for CD or related to intestinal inflammation in general. The association between the gene region containing *GLS* and other autoimmune traits, many of which show an increased risk of metabolic disease, could suggest that downregulation of IGN could also be present in other inflammatory diseases. Thus, further studies of IGN in other diseases are warranted. Also, the study does not answer if impaired IGN is involved in the risk of developing the disease or if it is a response to other disease-initiating mechanisms in CD. Moreover, while we see a significant correlation between decreased IGN gene expression and the degree of damage in the mucosa, as previously mentioned, the study does not answer if treatment with a gluten-free diet leading to healing of the intestinal mucosa restores expression of genes involved in IGN.

In conclusion, children with untreated CD show downregulation of genes critical for IGN in the intestinal mucosa, suggesting impairment of IGN. An impaired IGN may explain the increased risk of metabolic diseases like NAFLD found in CD patients as adults. Further studies around IGN and its role both in CD and other diseases are warranted.

## Data Availability

The data that support the findings of this study are available from the corresponding author, [ATN], upon reasonable request.

## Acknowledgements

We would like to extend our gratitude to all the families and patients who contributed to the study.

## Contributorship statement

OK and ÅTN conceived and carried out the experiments, performed data analysis and wrote the manuscript. HA, AHG and DA collected the biopsy material. Kaj Bjelkenkrantz, at the Department of Pathology, Unilabs, Stockholm, Sweden performed blinded review of all pathohistological diagnoses. AHG and HA revised the manuscript. DA suggested approaches for data analysis and revised the manuscript.

## Data availability statement

Datasets from the study are available from the corresponding author on reasonable request.

## Patient consent for publication

Not required.

## Patient and public involvement

Patients and/or the public did not take part in planning, performing, or reporting this research.

---

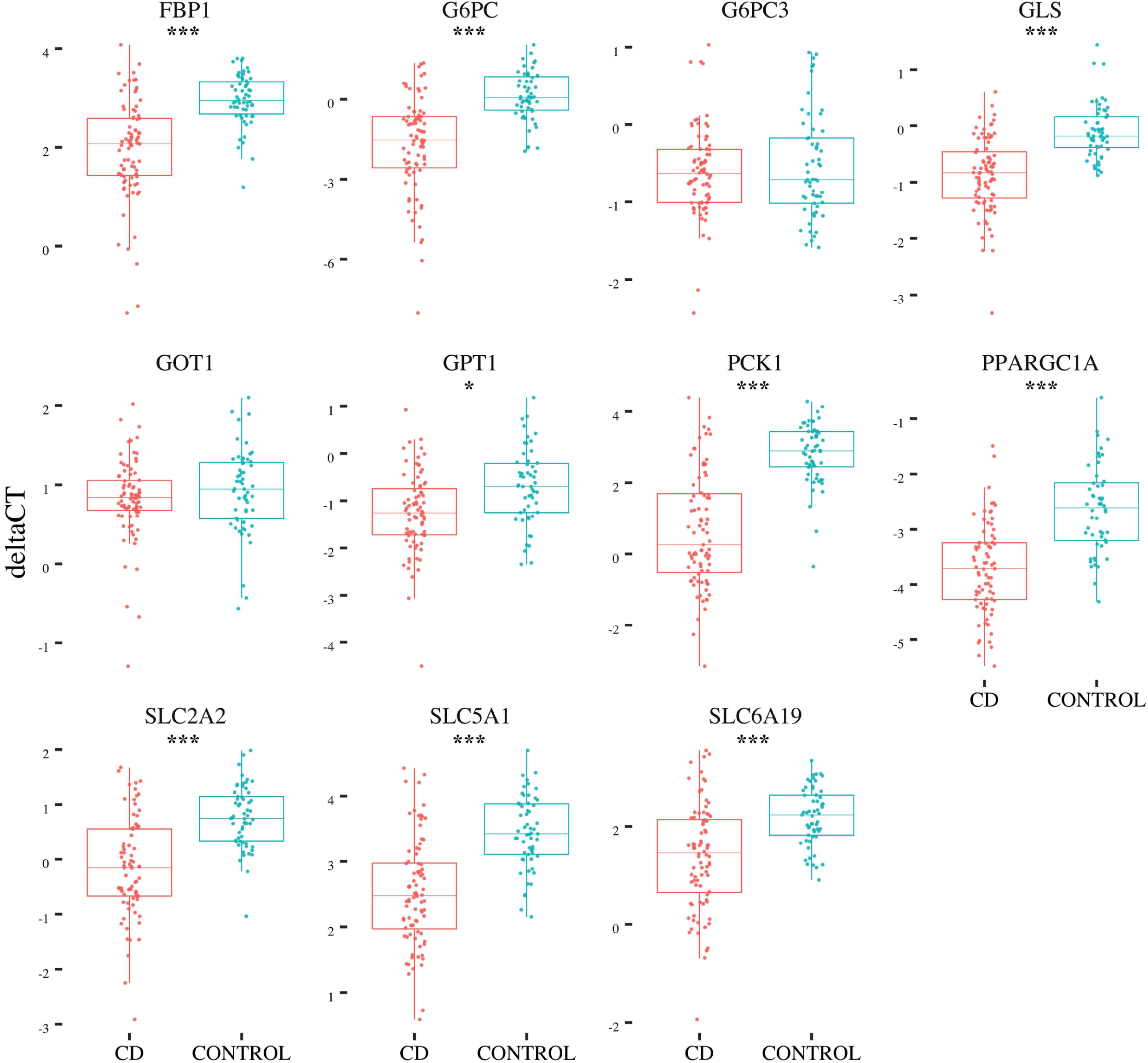

