## Supplemental Table 1 for "INTESTINAL GLUCONEOGENESIS IS DOWNREGULATED IN PAEDIATRIC PATIENTS WITH COELIAC DISEASE"

| **Supp. table 1.** Traits listed in the GWAS Catalog for the GLS/STAT1/STAT4 region (https://www.ebi.ac.uk/gwas) and the most associated SNP reported for each trait including their reported eQTLs* in the Genotype-Tissue Expression (GTEx) portal (https://www.gtexportal.org). | | | | | | | | |
| --- | --- | --- | --- | --- | --- | --- | --- | --- |
| **Trait** | **Study accession** | **Variant and risk allele** | **Chr** | **Location (bp)** | **P-value** | **Mapped gene** | **eQTL gene** | **tissue** |
| Systemic lupus erythematosus (SLE) | GCST011956 | rs11889341-T, rs7568275-? | 2 | 191079016 | 5.00E-123 | STAT4 | GLS | Colon - Transverse |
| Oral ulcer | GCST007839 | rs11684030-A | 2 | 191152153 | 1.00E-42 | STAT4, HMGB1P27 | STAT4 | Esophagus - Muscularis |
| Systemic scleroderma, rheumatoid arthritis, myositis, SLE | GCST007278 | rs10174238-? | 2 | 191108308 | 3.00E-42 | STAT4 |  |  |
| Thyroid preparation use measurement | GCST007932 | rs11889341-T | 2 | 191079016 | 3.00E-39 | STAT4 |  |  |
| Hypothyroidism | GCST007073 | rs7582694-? | 2 | 191105394 | 3.00E-39 | STAT4 |  |  |
| Autoimmune thyroid disease | GCST010571 | rs7568275-G | 2 | 191101726 | 5.00E-39 | STAT4 | GLS | Colon - Transverse |
| Mean platelet volume | GCST90002346 | rs1921911-C | 2 | 190855644 | 6.00E-31 | GLS | GLS | Muscle - Skeletal |
| Rheumatoid arthritis | GCST90013534 | rs11889341-T | 2 | 191079016 | 4.00E-30 | STAT4 |  |  |
| Reticulocyte count | GCST90002385 | rs398105120-AT | 2 | 190944197 | 4.00E-25 | STAT1, GLS |  |  |
| Primary biliary cirrhosis | GCST005581 | rs7574865-C, rs3024921-T | 2 | 191099907 | 9.00E-25 | STAT4 |  |  |
| Reticulocyte measurement | GCST90002387 | rs11687659-A | 2 | 190876471 | 6.00E-24 | GLS | GLS | Muscle - Skeletal |
| Autoimmune disease | GCST007071 | rs7568275-? | 2 | 191101726 | 2.00E-24 | STAT4 | GLS | Colon - Transverse |
| Body height | GCST007841 | rs35917062-? | 2 | 190796102 | 9.00E-23 | GLS |  |  |
| Systemic scleroderma | GCST009131 | rs3821236-A | 2 | 191038032 | 2.00E-23 | STAT4 |  |  |
| Juvenile idiopathic arthritis | GCST90010715 | rs11889341-T | 2 | 191079016 | 2.00E-19 | STAT4 |  |  |
| Sjogren syndrome | GCST002217 | rs10168266-T | 2 | 191071078 | 2.00E-17 | STAT4 |  |  |
| Inflammatory bowel disease | GCST003043 | rs1517352-C | 2 | 191066738 | 4.00E-14 | STAT4 |  |  |
| Systemic, polyarticular and oligoarticular juvenile idiopathic arthritis, rheumatoid factor negative | GCST005528 | rs10174238-G | 2 | 191108308 | 1.00E-13 | STAT4 |  |  |
| Multiple sclerosis | GCST009597 | rs6738544-C | 2 | 191124630 | 2.00E-13 | STAT4 |  |  |
| Limited scleroderma | GCST005555 | rs10174238-? | 2 | 191108308 | 1.00E-11 | STAT4 |  |  |
| Systemic scleroderma, SLE | GCST002069 | rs7601754-? | 2 | 191075725 | 3.00E-11 | STAT4 |  |  |
| Rheumatoid arthritis, celiac disease | GCST008644 | rs6749371-? | 2 | 191037458 | 7.00E-11 | STAT4 |  |  |
| PHF-tau measurement | GCST010340 | rs7033192-? x rs6434425-? | 2 | 190857590 | 6.00E-10 | SLC8A1 - GLS |  |  |
| Birth weight | GCST009437 | rs1547550-C | 2 | 190980999 | 1.00E-10 | STAT1 | GLS/STAT4 | Muscle - Skeletal/Nerve - Tibial |
| Mean corpuscular hemoglobin | GCST007068 | rs7565237-? | 2 | 190971964 | 3.00E-10 | STAT1 | GLS | Muscle - Skeletal |
| Crohn's disease | GCST003044 | rs1517352-C | 2 | 191066738 | 1.00E-10 | STAT4 |  |  |
| Hepatocellular carcinoma | GCST001775 | rs7574865-G | 2 | 191099907 | 2.00E-10 | STAT4 |  |  |
| Immune system disease | GCST000987 | rs7574865-T | 2 | 191099907 | 4.00E-10 | STAT4 |  |  |
| Non-typhoidal Salmonella bacteremia | GCST005590 | rs13390936-T | 2 | 191090090 | 9.00E-10 | STAT4 |  |  |
| BMI-adjusted waist-hip ratio | GCST009858 | rs78219026-? | 2 | 190881507 | 3.00E-09 | GLS | GLS | Artery - Aorta |
| Waist-hip ratio | GCST007067 | rs6434426-? | 2 | 190861883 | 7.00E-09 | GLS | GLS | Artery - Tibial |
| Neutrophil percentage of granulocytes | GCST004623 | rs60976990-T | 2 | 190991701 | 2.00E-09 | STAT1 |  |  |
| Behcet's syndrome | GCST001804 | rs7574070-A | 2 | 191145762 | 1.00E-09 | STAT4 | STAT4 | Cells - EBV-transformed lymphocytes |
| Anti-centromere-antibody-positive systemic scleroderma | GCST005552 | rs10174238-? | 2 | 191108308 | 2.00E-09 | STAT4 |  |  |
| Ulcerative colitis | GCST003045 | rs1517352-C | 2 | 191066738 | 2.00E-09 | STAT4 |  |  |
| Eosinophil count | GCST90002381 | rs6752770-G | 2 | 191108837 | 2.00E-09 | STAT4 |  |  |
| Type I diabetes mellitus | GCST90013445 | rs7582694-G | 2 | 191105394 | 3.00E-09 | STAT4 |  |  |
| Celiac disease | GCST009874 | rs6749371-? | 2 | 191037458 | 7.00E-09 | STAT4 |  |  |
| Self reported educational attainment | GCST006442 | rs66721975-A | 2 | 190836459 | 2.00E-08 | GPR39, GLS |  |  |
| Protein measurement | GCST011427 | rs2066799-T | 2 | 190986759 | 3.00E-08 | STAT1 |  |  |
| JT interval, response to sulfonylurea | GCST004032 | rs12468579-? | 2 | 190967538 | 5.00E-08 | STAT1 | GLS | Muscle - Skeletal |
| Amyloid-beta measurement | GCST010339 | rs12065191-? x rs3024861-? | 2 | 191059880 | 8.00E-08 | STAT4 |  |  |
| Rheumatoid arthritis, Crohn's disease | GCST90016610 | rs12612769-? | 2 | 191089272 | 2.00E-07 | STAT4 |  |  |
| Rheumatoid arthritis, ulcerative colitis | GCST90016625 | rs11889341-? | 2 | 191079016 | 2.00E-07 | STAT4 |  |  |
| Vitiligo | GCST004785 | rs199559999-? | 2 | 191090480 | 3.00E-07 | STAT4 |  |  |
| Longevity | GCST009448 | rs13033350-A | 2 | 190826756 | 1.00E-06 | GLS | GLS | Muscle - Skeletal |
| Diabetes mellitus, coronary artery disease | GCST90014128 | rs148894474-C | 2 | 190815604 | 3.00E-06 | GLS |  |  |
| Threonine measurement | GCST009391 | rs1607187-? | 2 | 190950312 | 3.00E-06 | GLS, STAT1 | GLS | Muscle - Skeletal |
| Biliary liver cirrhosis | GCST001685 | rs7574865-T | 2 | 191099907 | 1.00E-06 | STAT4 |  |  |
| Myositis | GCST006051 | rs4853540-? | 2 | 191052591 | 2.00E-06 | STAT4 |  |  |
| *eQTL = expression Quantitative Trait |  |  |  |  |  |  |  |  |
